# The Histopathological Diagnosis of Adenocarcinoma & Squamous Cells Carcinoma of Lungs by Artificial intelligence: A comparative study of convolutional neural networks

**DOI:** 10.1101/2020.05.02.20044602

**Authors:** Mohammad Ali Abbas, Syed Usama Khalid Bukhari, Asmara Syed, Syed Sajid Hussain Shah

**Author notes:** Corresponding Author: 1. Syed Usama Khalid Bukhari, a., b. ORCID ID: https://orcid.org/0000-0003-1581-8609, c. Assistant Professor, Department of Computer Science, The University of Lahore, Islamabad, Pakistan. Student (MCSC), Department of Computer Science, The University of Lahore, Islamabad, Pakistan. Assistant Professor, Department of Computer Science, The University of Lahore, Islamabad, Pakistan. Lecturer, Faculty of Medicine, Northern Border University, Arar-Kingdom of Saudi Arabia. Professor, Faculty of Medicine, Northern Border University, Arar-Kingdom of Saudi Arabia.

## Abstract

**Introduction:** Malignant tumors of the lung are the most important cause of morbidity and mortality due to cancer all over the world. A rising trend in the incidence of lung cancer has been observed. Histopathological diagnosis of lung cancer is a vital component of patient care. The use of artificial intelligence in the histopathological diagnosis of lung cancer may be a very useful technology in the near future.

**Aim:** The aim of the present research project is to determine the effectiveness of convolutional neural networks for the diagnosis of squamous cell carcinoma and adenocarcinoma of the lung by evaluating the digital pathology images of these cancers.

**Materials & Methods:** A total of 15000 digital images of histopathological slides were acquired from the LC2500 dataset. The digital pathology images from lungs are comprised of three classes; class I contains 5000 images of benign lung tissue, class II contains 5,000 images of squamous cell carcinoma of lungs while Class III contains 5,000 images of adenocarcinoma of lungs. Six state of the art off the shelf convolutional neural network architectures, VGG-19, Alex Net, ResNet: ResNet-18, ResNet-34, ResNet-50, and ResNet-101, are used to assess the data, in this comparison study. The dataset was divided into a train set, 55% of the entire data, validation set 20%, and 25% into the test data set.

**Results:** A number of off the shelf pre-trained (on ImageNet data set) convolutional neural networks are used to classify the histopathological slides into three classes, benign lung tissue, squamous cell carcinoma-lung and adenocarcinoma - lung. The F-1 scores of AlexNet, VGG-19, ResNet-18, ResNet-34, ResNet-50 and ResNet-101, on the test dataset show the result of 0.973, 0.997, 0.986, 0.992, 0.999 and 0.999 respectively.

**Discussion:** The diagnostic accuracy of more 97% has been achieved for the diagnosis of squamous cell carcinoma and adenocarcinoma of the lungs in the present study. A similar finding has been reported in other studies for the diagnosis of metastasis of breast carcinoma in lymph nodes, basal cell carcinoma, and prostatic cancer.

**Conclusion:** The development of algorithms for the recognition of a specific pattern of the particular malignant tumor by analyzing the digital images will reduce the chance of human errors and improve the efficiency of the laboratory for the rapid and accurate diagnosis of cancer.

## Introduction

Lung cancer has become a global health issue [1]. The malignant tumors of the lung are the most common cause of death due to cancers in men and these neoplasms are the second most common cause of cancer-associated mortality in female all over the world [2]. The prevalence of lung carcinomas is associated with age, gender, ethnicity, socioeconomic conditions, and regions with their smoking patterns. The incidence of lung cancer is increasing which has been attributed to different factors such as worldwide higher prevalence of tuberculosis, the diesel exhaust exposure (elemental carbon) in certain occupations, rise in the number of aged people (age 60 years or above) in the community [3–5]. Smoking is more associated with small cell carcinoma and squamous cell carcinoma of the lungs [6]. Lung cancer is most prevalent in the elderly population. Most of the cases of lung cancer are diagnosed in age between 40 to 90 years with the mean age of 65 years [7]. Squamous cell carcinoma and adenocarcinoma are the most prevalent types of lung cancer while the other histological types include small cell carcinoma and large cell carcinomas. The mortality and morbidity of lung cancer are associated with the stage and distant recurrence of the tumors. The enlargement of the hilar lymph node (more than one centimeter) is an important predictor of the distant recurrence [8].

The rising trend of lung cancer in the world is going to further increase the morbidity and mortality associated with malignancy. The effective management of malignant tumors of lungs is required for the reduction in the rate of morbidity and mortality. In this regard, the accurate and rapid diagnosis of pulmonary tumors is of vital importance. Histopathological examination is required for the conclusive diagnosis of malignant tumors of lungs which is based on the morphological features of cells and patterns of arrangement of these neoplastic cells. The increase in the incidence of lung cancer will raise the burden on health professionals including the histopathologists who are involved in the diagnosis. This raises an urgent need for the establishment of a computer-aided diagnostic system that could help in the diagnosis of pulmonary malignant tumors.

With the development of digital scanners for the whole slide, it becomes quite possible to have the high-resolution digital images of the hematoxylin and eosin-stained slides of tissue sections from different pathological lesions. The application of algorithms for the image analysis of digital pathology images revealed encouraging results but there were certain limitations in the analysis of images of pathological lesions. The limitations were overcome by the development of deep machine learning technology. The aim and objectives of this research project are to find out the effectiveness of convolutional neural networks (CNN) for the diagnosis of common types of lung carcinoma (squamous cell carcinoma and adenocarcinoma) by the evaluation of digital images of these cancers.

### Materials & Methods

A comparison study is conducted, where three CNN architectures and four variations of them are used to show, how off the shelf CNN architectures perform on digital histopathological slides for a classification problem. A total of fifteen thousand (15000) digital images of lung tissues are used to classify three classes in the data set. Class I, is composed of five thousand (5000) digital images of histopathological slides labeled as benign lung tissue. Class II, is also composed of five thousand (5000) digital images of histopathological slides labeled as of lung squamous cell carcinoma. Class III, is also composed of five thousand (5000) digital images of histopathological slides labeled as lung adenocarcinoma. The dataset is acquired from Lung and Colon Cancer Histopathological Image Dataset (LZ2500). The images are categorized, labeled, cleaned, and augmented by the same group [9].

For the assessment, the data is divided into three sets Test set, Train set, and Validation set. The Train set contains two thousand seven hundred and fifty (2750) images from every class of data, so in total it contains eight thousand two hundred and fifty (8250) images, which is 55% of the total data set. The test set contains twelve hundred and fifty (1248) images from every class of data, so in total it contains three thousand seven hundred and forty-four (3744) images, which is 25% of the total data set. Where the validation set contains approximately one thousand images (998–1002) images from every class of data, so in total it contains approximately three thousand (2994–3006) images, which is 20% of the total data set. The data is randomly put in the respected sets using the FAST-AI library. We applied augmentation on the data set. Detail of the parameters for architectures and augmentation is present in table 1.

**Table 1:** Parameters for Image augmentations

| *Type of Augmentation* | *Description* |
| --- | --- |
| Horizontal & Vertical flip | Image is flipped both horizontally and vertically with the 0.5 probability |
| random zoom | Image is zoomed range = 0 to 10% with 75% probability. |
| random lighting | Change in the light (brightness) of the image range = 0 to 40% with 75% probability. |
| Image Size | 224 x 224 |

All the architectures were loaded with pre-trained weights trained on ImageNet [10]. Same hyperparameters along with the customized last fully connected layer added at the end of them. The configuration of the last layer is presented in table 2.

**Table 2:** Last layer configuration

| *Layer* | *Description* |
| --- | --- |
| *DropOut* | With rate = 0.3 |
| *Linear Layer* | Input: number of features from Arch Pooling Layer, output: 128 |
| *DropOut* | With rate = 0.5 |
| *Linear Layer* | Input: 128, output: 3 |

Hyper parameters used are

- learning rate: 1e-5
- Momentum: 0.1
- Batch Size: 32

## Results

A total of fifteen thousand (15000) digital images have been analyzed which are divided into three categories of class I, classII and class III. The class I is comprised of 5000 images of benign lung tissue, class II is contains 5000 images of squamous cell carcinoma of lungs and there are 5000 images of adenocarcinoma of lungs in the category of class III. Four ResNet architecture models, ResNet-18, ResNet-34 ResNet-50, ResNet 101, VGG 19 and Alex-Net are re used to classify the input image. Accuracy of 98.8%,99.18 %, 99.6,99.8 %, 98.93% and 97.26% % and f1-score of 98.64, 99.28,99.95,99.98,99.75 and 97.38 respectively was obtained on the unseen test data set. The results are shown in table 3 and the classification results in the form of the confusion matrix and ROC curves for test data are depicted in Figure 1.

**Table 3:** Result of all architectures

| Arch | Accuracy | Precision | Recall | F-1 Score |
| --- | --- | --- | --- | --- |
| **Alex-Net** | 97.26% | 0.9767628205 | 0.971 | 0.9738728851 |
| **ResNet-18** | 98.8% | 0.9930555556 | 0.98 | 0.986484584 |
| **ResNet-34** | 99.18% | 0.9957264957 | 0.99 | 0.9928549907 |
| **ResNet-50** | 99.6% | 0.9991987179 | 1 | 0.9995991984 |
| **ResNet-101** | 99.8% | 0.999732906 | 1 | 0.9998664352 |
| **VGG 19** | 98.93% | 0.9951923077 | 1 | 0.9975903614 |

**Figure 1.**
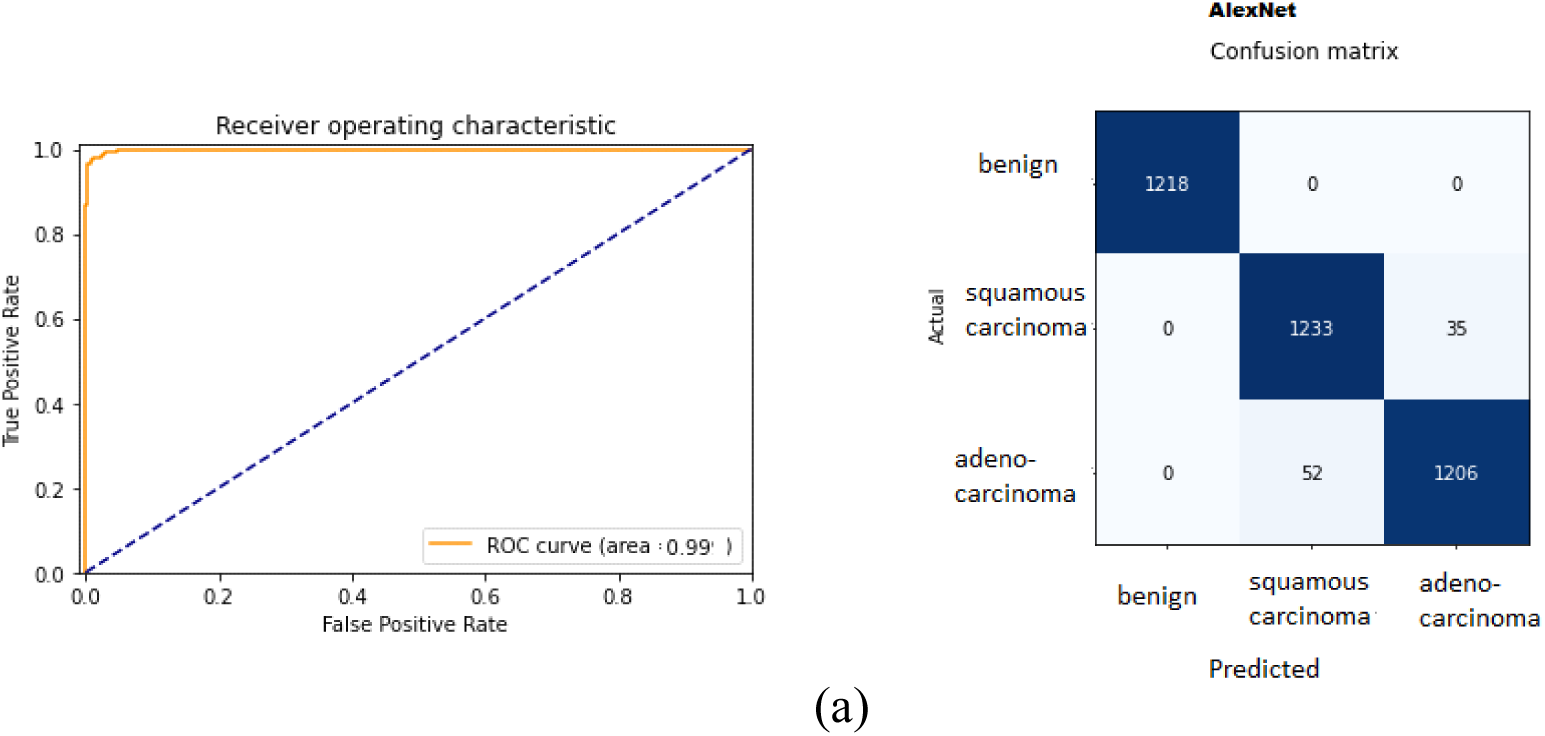
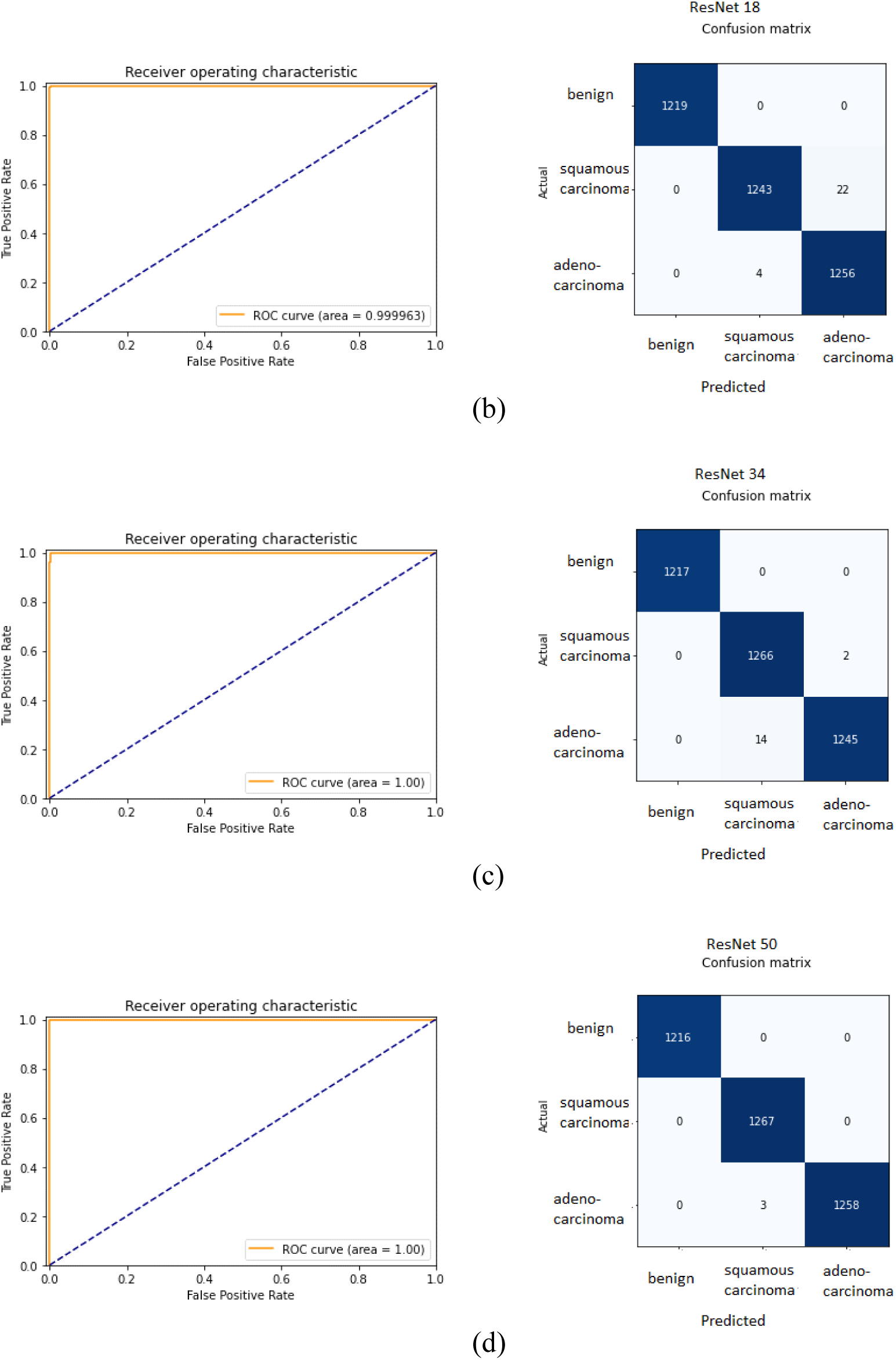
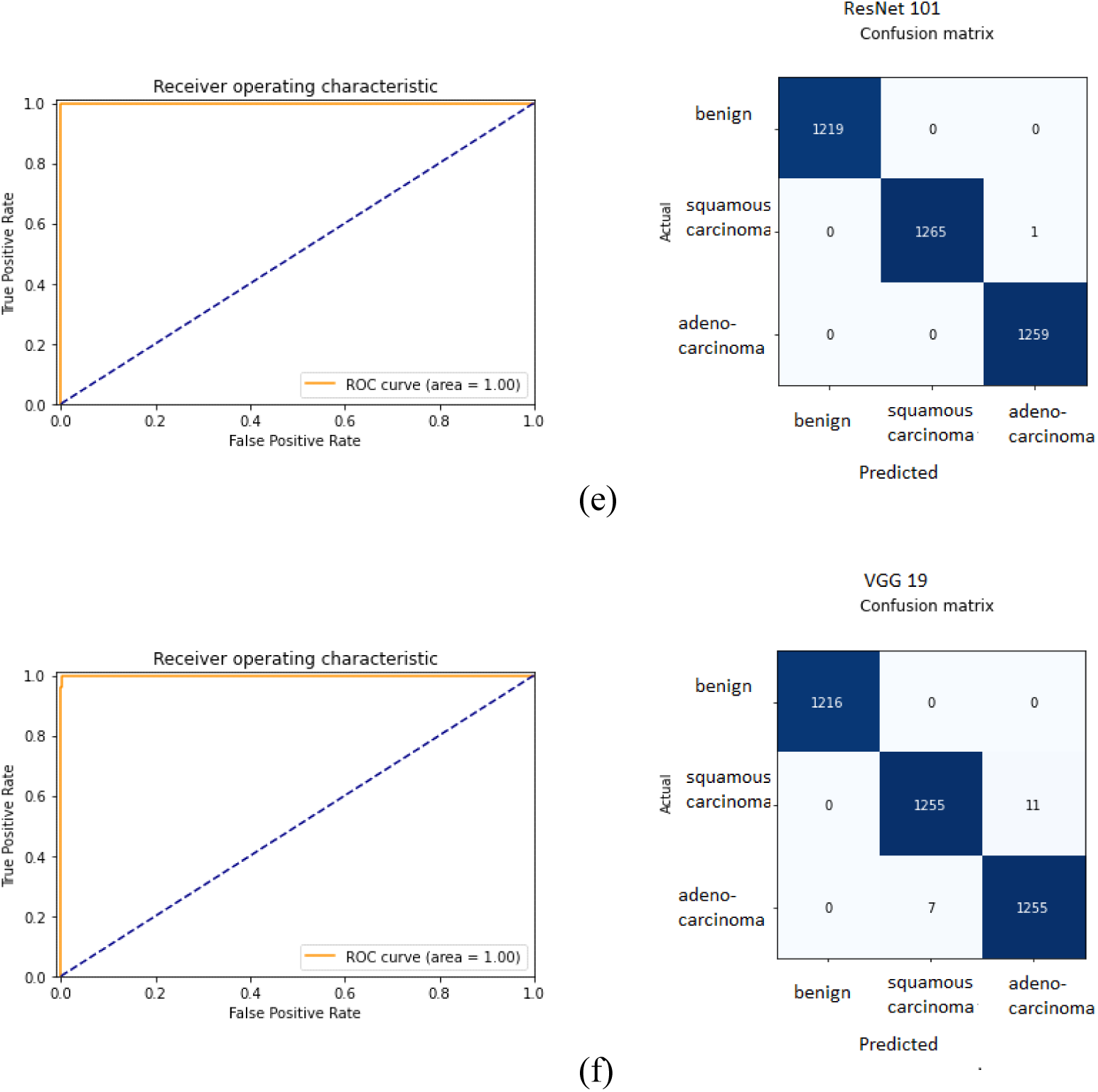
showing the ROC curve and confusion matrix of the CNN architectures (a) AlexNet (b) ResNet 18 (c) ResNet 34 (d) ResNet 50 (e) ResNet 101 (f) VGG-19.

**Figure 2:**
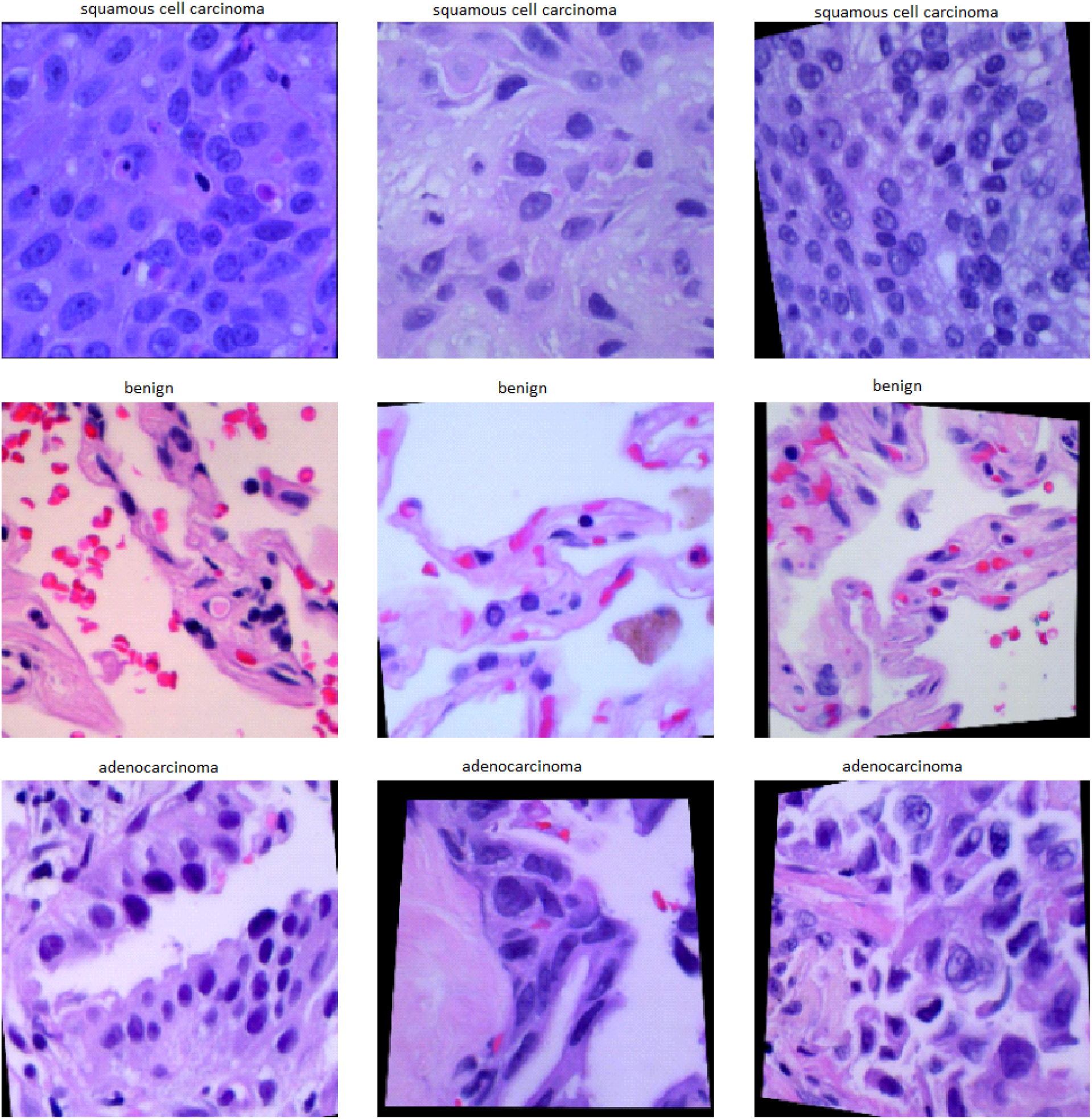
Data bunch generated by FastAI API (Augmented) to show it loaded the images and label correctly,

## Discussion

The use of a computer with the application of artificial intelligence to analyze the data for the extraction of useful and important information could be a very helpful tool in diagnostics.

The machine learning-based computer programs can be used to find out the solutions by evaluating and analyzing the complex data in the field of medicine which may improve patient care. The application of artificial intelligence in medicine could be helpful in improving patient care by reducing the chance of error and improving the speed of delivery of desirable health care support with cost-effectiveness. The development of a computerized system for the treatment of patients has revealed a reduction in medication errors [11].

The laboratory diagnosis has got paramount importance in the management of the disease. The significant number of decisions in medicine depends upon the report of laboratory tests. The use of a computer-aided program could assist the pathologist in improving the speed and reducing the error rate in the laboratory diagnosis. Histopathology is a branch of pathology which deals with the examination of a tissue specimen for the diagnosis which is based on the structural changes along with the formation of specific patterns by the cells in the morbid tissue. The computer vision-based system may be a valuable tool in the assessment of biopsy specimens for the histopathological diagnosis. The development of algorithms for the recognition specific pattern of the particular lesions by analyzing the digital images will reduce the stress on the histopathologist and improve the efficiency of the laboratory.

The development of digital analysis of the images is expected to play an important role in improving the precision of histomorphological assessment as the deep machine learning yielded advancement in the computational pathology [12]. There are many types of deep neural networks. In computational pathology for the analysis of microscopic digital slides of the pathological lesions, the convolutional neural network is most frequently adopted for the image analysis [13]. The convolutional neural network (CNN) is characterized by an input layer, many hidden layers, and an output layer [14].

In the present study, CNN architectures are used to analyze fifteen thousand (15000) digital images of lung tissues which included five thousand (5000) digital images of lung adenocarcinoma, five thousand (5000) digital images of lung squamous cell carcinoma and five thousand (5000) digital images of benign lung tissue and the diagnostic accuracy is over 97%. The results are in accordance with the other studies which were carried out for detection of metastasis of breast carcinoma in lymph nodes, basal cell carcinoma, prostatic cancer, and non-small cell cancers of lungs [15–17].

These encouraging results of the application of deep machine learning methodologies in the field of histopathology are predicting the rising sun of artificial intelligence-based computer systems for the cancer diagnosis in the near future. These computer-aided systems for the histopathological diagnosis will improve patient care by rapid and accurate diagnosis with increased reproducibility by reducing the inter-observer variation. The use of artificial intelligence in the fields of histopathology may also reduce the chance of human errors which may occur due to the fatigability of eyes and brain in human beings. In this emerging field of artificial intelligence in diagnostic histopathology, the histopathologists are urged to update and equip themselves with the computer vision-based programs so that this technology may be evaluated more deeply and critically which is vital for further improvement in the technology for better accuracy and efficiency. Further studies of convolutional neural network (CNN) on different histopathological types and subtypes of lesions of human tissue are recommended.

## Conclusion

The development of algorithms for the recognition of a specific pattern of the particular lesions by analyzing the digital images will reduce the stress on the histopathologists leading to less chance of human error and improve the efficiency of the laboratory for the rapid and accurate diagnosis of malignant tumors

## Data Availability

Data is provided by 
Borkowski AA, Bui MM, Thomas LB, Wilson CP, DeLand LA, Mastorides SM, and made it available for all researchers.  Rhe Lung and Colon Cancer Histopathological Image Dataset (LC25000). 2019. arXiv:1912.12142

## Acknowledgement

The authors are thankful to Borkowski AA, Bui MM, Thomas LB, Wilson CP, DeLand LA, Mastorides SM for making available the Lung *and Colon Cancer Histopathological Image Dataset (LC25000). 2019. arXiv:1912.12142* for AI research. The authors are also thankful to DockCloud company for authorizing the authors to use of their hardware and services for the purpose of this study.

## References

1. Mao Y, Yang D, He J, Krasna MJ. Epidemiology of Lung Cancer. Surg Oncol Clin N Am. 2016;25(3):439–45. doi: 10.1016/j.soc.2016.02.001.

2. Torre LA, Siegel RL, Jemal A. Lung Cancer Statistics. Adv Exp Med Bio. 2016;893;1–19. doi: 10.1007/978-3-319-24223-1_1.

3. Hong S, Mok Y, Jeon C, Jee SH, Samet JM. Tuberculosis, smoking and risk for lung cancer incidence and mortality. Int J Cancer. 2016: 139(11):2447–55. doi: 10.1002/ijc.30384.

4. Ge C, Peters S, Olssona, Portengen L, Schuz J, Almansa J et al. Diesel Engine Exhaust Exposure, Smoking, and Lung Cancer Subtype Risks: A Pooled Exposure-response Analysis of 14 Case-control Studies. Am J Respire Crit Care Med. 2020 Apr 24. doi: 10.1164/rccm.201911-2101OC

5. Donnelly D W, Anderson LA, Gavin A. Cancer incidence projections in Northern Ireland to 2040. Cancer Epidemiol Biomarkers Prev. 2020 Apr 24. pii: cebp.0098.2020. doi: 10.1158/1055-9965.EPI-20-0098.

6. Zia N, Khokhar MA, Tabassum MN, Qamar S, Tirmizi IQ, Qudratullah M, et al. Trends in Development of Various Types of Lung Cancer after Cessation of Smoking. P J M H S. 2017: 11(1); 140–44

7. Davoudadham, Moradi-Asl E, Abbasgholizade N, Zandian H, Abazari M, Ghobadi H. Spatial Analysis and Epidemiology of Lung Cancer in the Northwest of Iran. P J M H S. 2018:12(4);1851–55.

8. Duran FM, Akın SE, Calik M, Esme H. The factors affecting the distant recurrence in patients with non-small cell lung cancer (NSCLC) and our results. Ann Clin Anal Med 2020;11(1):28–32

9. Borkowski AA, Bui MM, Thomas LB, Wilson CP, DeLand LA, Mastorides SM. Lung and Colon Cancer Histopathological Image Dataset (LC25000). 2019. arXiv:1912.12142

10. Russakovsky O, Deng J, Su H, Krause J, Satheesh S, Ma S et al. Imagenet large scale visual recognition challenge. International journal of computer vision. 2015:115(3); 211–252. doi.org/10.1007/s11263-015-0816-y

11. Jia P, Zhang L, Chen J, Zhao P, Zhang M. The Effects of Clinical Decision Support Systems on Medication Safety: An Overview. PLoS One. 2016;11(12):e0167683. doi:10.1371/journal.pone.0167683

12. Acs B, Rantalainen M, Hartman J. Artificial intelligence as the next step towards precision pathology. J Intern Med; 2020;https://doi.org/10.1111/joim.13030

13. Chang HY, Jung CK, Woo JI, Lee S, Cho J, Kim SW, et al. Artificial Intelligence in Pathology. J Pathol Transl Med. 2019;53(1):1–12. doi: 10.4132/jptm.2018.12.16.

14. Janowczyk A, Madabhushi A. Deep learning for digital pathology image analysis: A comprehensive tutorial with selected use cases. J Pathol Inform. 2016;7:29. doi:10.4103/2153-3539.186902

15. Ehteshami Bejnordi B, Veta M, Johannes van Diest P, van Ginneken B, Karssemeijer N, Litjens G et al. Diagnostic assessment of deep learning algorithms for detection of lymph node metastases in women with breast cancer. JAMA 2017;318(22):2199–2210. doi: 10.1001/jama.2017.14585

16. Campanella G, Hanna MG, Geneslaw L, Miraflor A, Silva VWK, Busam KJ et al. Clinical-grade computational pathology using weakly supervised deep learning on whole slide images. Nat Med 2019; 25: 1301–1309. doi: 10.1038/s41591-019-0508-1.

17. Coudray N, Ocampo PS, Sakellaropoulos T, Narula N, Snuderl M, Fenyo D et al. Classification and mutation prediction from non-small cell lung cancer histopathology images using deep learning. Nat Med 2018; 24: 1559–67. doi: 10.1038/s41591-018-0177-5.

